# Balancing Equity and HLA Matching in Deceased-Donor Kidney Allocation with Eplet Mismatch

**DOI:** 10.1101/2024.06.13.23290644

**Authors:** Michal A. Mankowski, Loren Gragert, Dorry L. Segev, Robert Montgomery, Sommer E. Gentry, Massimo Mangiola

**Affiliations:** Transplant Institute, NYU Langone Health, New York, New York; Department of Surgery, NYU Grossman School of Medicine, NYU Langone Health, New York, NY, USA; Division of Biomedical Informatics and Genomics, Deming Department of Medicine, Tulane University School of Medicine, New Orleans, LA, USA; Department of Population Health, NYU Grossman School of Medicine, New York, NY, USA

**Keywords:** allocation, histocompatibility, eplet mismatch, simulation

## Abstract

**Background:** Prioritization of HLA antigen-level matching in the US kidney allocation system intends to improve post-transplant survival but causes racial disparities and thus has been substantially de-emphasized. Recently, molecular matching based on eplets has been found to improve risk stratification compared to antigen matching.

**Methods:** To assign eplets unambiguously, we utilized a cohort of 5193 individuals with high resolution allele-level HLA genotypes from the National Kidney Registry. Using repeated random sampling to simulate donor-recipient genotype pairings based on the ethnic composition of the historical US deceased donor pool, we profiled the percentage of well-matched donors for candidates by ethnicity.

**Results:** The percentage of well-matched donors with zero-DR/DQ eplet mismatch was 3-fold less racially disparate for Black and Asian candidates than percentage of donors with zero-ABDR antigen mismatches, and 2-fold less racially disparate for Latino candidates. For other HLA antigen and eplet mismatch thresholds, the percentage of well-matched donors was more similar across candidate ethnic groups.

**Conclusions:** Compared to the current zero-ABDR antigen mismatch, prioritizing a zero-DR/DQ eplet mismatch in allocation would decrease racial disparities and increase the percentage of well-matched donors. High resolution HLA deceased donor genotyping would enable unambiguous assignment of eplets to operationalize molecular mismatch metrics in allocation.

**Key Points:** *Question:* What is the impact of prioritizing low molecular mismatch transplants on racial and ethnic disparities in US deceased-donor kidney allocation, compared to the current prioritization of antigen-level matching?

*Findings:* The lowest-risk eplet mismatch approach decreases racial disparities up to 3-fold compared to lowest-risk antigen mismatch and identifies a larger number of the lowest allo-immune risk donors.

*Meaning:* Prioritizing eplet matching in kidney transplant allocation could both improve outcomes and reduce racial disparities compared to the current antigen matching.

## 1. INTRODUCTION

Closer donor-recipient HLA matching is associated with improved post-transplant kidney graft survival.^1,2^ However, prioritizing HLA matching comes with a tradeoff of increased transplant rates for candidates with more frequent HLA genotypes, which disadvantages minority populations who carry greater HLA diversity. Points for HLA-B matching were removed from the deceased donor kidney allocation because HLA-B matching increased racial disparities while correlating with graft survival only slightly.^3–5^ At present, only antigen-level zero-ABDR, zero-DR, and 1-DR mismatches are awarded points in the US deceased donor kidney allocation. A prior study of the OPTN database of recovered donors and waitlist registrations found that for the lowest-risk category of highly prioritized zero-ABDR antigen mismatches, White candidates have 6 times as many 0-ABDR mismatched donors as African Americans have, and 9 times as many as Asians have^6^

A redesign of allocation policy for “continuous distribution” is under development and plans to include priority points only for zero- and 1-DR antigen mismatches, as a new outcomes study by the Scientific Registry for Transplant Recipients (SRTR) found zero-DR and zero-ABDR antigen mismatch had similar survival and there was no independent impact of HLA-A and -B antigen matching.^9^ The American Society of Transplant Surgeons (ASTS), in its public comment on the kidney/pancreas continuous distribution concept paper, recognized the benefits of HLA-DR matching but simultaneously raised concerns about its fairness for minority populations.^7^ These concerns could be addressed by identifying strategies to mitigate disparities while maintaining the survival benefits of HLA matching.^8^

Advances in genotyping technology, such as Next Generation Sequencing (NGS), have enabled allele-level HLA compatibility assessment between donors and recipients.^9^ Currently, reporting of antigen-level typing is the standard practice, as unambiguous allele-level information is not usually available for deceased donor kidney allocation. Antigen-level typing broadly categorizes HLA molecules based on serological reactivity, concealing some clinically-relevant differences between HLA proteins. Allele-level typing identifies the specific gene sequences, enabling eplet mismatch to be directly computed from the HLA amino acid sequence of the alleles. Rapid long-read NGS technology being tested in clinical HLA laboratories could soon provide allele-level genotyping of deceased donors routinely at the time of allocation.^10^

Eplet matching promises improved transplant outcomes by providing a more precise assessment of donor-recipient compatibility through examining critical amino acid motifs on HLA proteins that are predicted to influence specificity of anti-HLA antibody binding. Several studies have demonstrated that higher levels of HLA-DR and DQ eplet mismatches correlate with the formation of post-transplant de-novo Donor Specific Antibody (dnDSA), which is associated with antibody-mediated rejection^11–13^, and that eplet mismatch is a prognostic biomarker for both T cell (TCMR) and antibody-mediated rejection (ABMR).^14–16^

Because of the projected improvements in long term graft survival, the idea of utilizing eplet matching in kidney allocation is gaining community attention.^17–19^ Some living donation programs, such as National Kidney Registry (NKR)^20^ and Royal Children’s Hospital Melbourne kidney transplant program have already adopted eplet mismatch in their allocation systems.^21^

Detailed studies on equity and utility of molecular mismatch in the context of allocation are needed, as the Sensitization in Transplantation: Assessment of Risk (STAR) working group has advocated.^8^ The hope is that matching kidneys using HLA molecular mismatch strategies such as eplet mismatch would improve deceased donor kidney transplant outcomes while ensuring equity across racial and ethnic groups.^9^ To model equity in access among candidate ethnic groups to well-matched deceased donors in the US, we utilized a National Kidney Registry (NKR) cohort of over 5000 individuals with the high resolution HLA genotyping necessary to make unambiguous eplet assignments, as OPTN datasets lack the required HLA data.

## 2. METHODS

### 2.1 Study population

This study used HLA genotyping and ethnicity data from the National Kidney Registry (NKR), a nonprofit, 501© organization that facilitates kidney paired donations for members of its clinical network in the United States. The NKR dataset included 5193 recipients, and living donors that were all HLA genotyped at the allele-level enabling unambiguous assignment of eplets. Individual race/ethnicity (hereafter referred to more simply as “ethnicity”) was self-reported.

Twenty separate OPTN-representative donor pools of randomized 1000 individuals were sampled from the NKR cohort, each having the ethnic composition of the historical 5-year average from the OPTN deceased donor pool (Asian (2.5%), Black (14.5%), Hispanic/Latino (14.8%), White (66.8%), and Others (1.3%)) - see Table 1. After sampling each replicate donor pool, the percentage of well-matched donors was calculated for each of the remaining individuals (i.e., 4193 individuals from the NKR pool that were not in the OPTN-representative donor pool). Creating twenty resampled 1000 donors / 4193 candidates pools allowed us to capitalize on the larger sample size of the NKR dataset while adjusting for the ethnic composition, reducing the impact of sampling error. We aggregated the percentages of well-matched donors with individuals by their ethnicity.

**Table 1.**
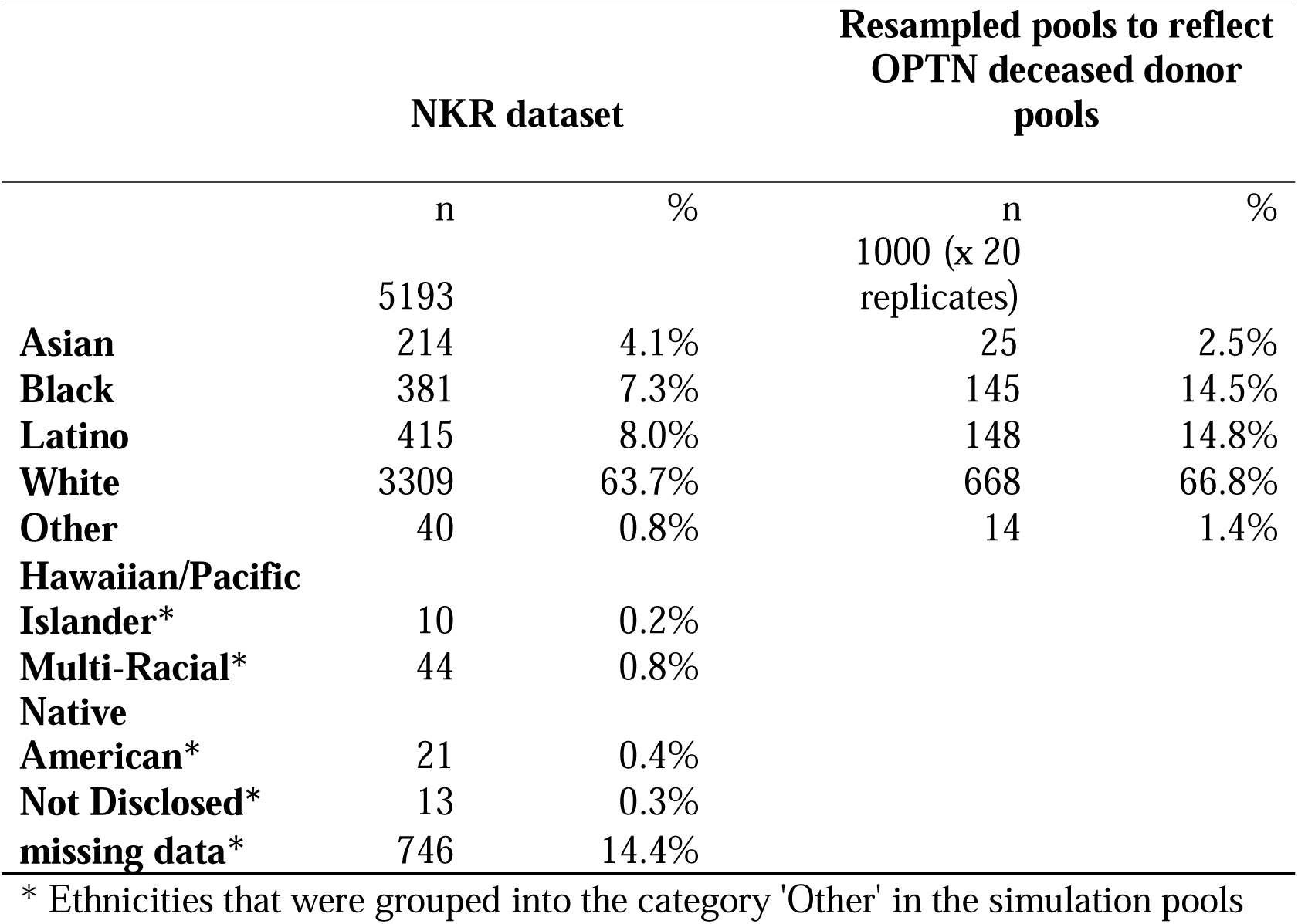
Study population. From a dataset of 5193 individuals with high-resolution HLA typing data from National Kidney Registries (NKR), we randomly sampled 20 replicate donor pools of 1000 individuals, each with pool reflecting the ethnic composition of the historical US deceased-donor pool. Ethnic distributions of the NKR dataset and the simulated donor pools are provided.

### 2.2 HLA genotyping and HLA eplet mismatch load

All 5193 individuals were fully genotyped at the allele-level for all 11 classical HLA loci (HLA-A, B, C, DRB1, DRB3/4/5, DQA1, DQB1, DPA1, DPB1). To assign antigen mismatch, each allele was mapped to corresponding HLA antigens according to the OPTN Histocompatibility tables and guidance.^22^ For assigning eplet mismatch, we used the publicly available calculator from the NKR,^23^ which is based on HLAMatchmaker, and which has been verified against the calculator in the HLA Eplet Registry^24^ yielding 100 percent concordance. For each candidate-donor pair, we calculated (1) the sum of eplet mismatches for HLA-DR (HLA-DRB1/3/4/5) and HLA-DQ (HLA-DQA1/HLA-DQB1) (2) and the number of A, B, DR, or DQ antigen mismatches. We defined a low risk eplet mismatch as a 1-10 DR mismatch and/or 1-10 DQ mismatch.

### 2.3 Simulation and Statistical Analysis

For each OPTN-representative donor pool, we calculated the percentage of well-matched donors for each candidate (also from NKR) in a resampled pool with the ethnic composition as the OPTN deceased donor pool, using either the antigen mismatch (aMM) or eplet mismatch (epMM) risk categories. The percentage of well-matched donors is the percentage of donors that have a given HLA mismatch level with a candidate. In our main analyses, candidates were matched solely based on HLA compatibility, not accounting for ABO blood group compatibility or any other allocation criteria.

The cohort resampling was programmed in Python 3.9 and the package Scipy 1.9.2 was used for statistical calculations. For each of the 20 simulation runs, the ratios of the percentage of well-matched donors between candidate ethnicities were calculated to obtain a 95% confidence interval (CI).

## 3. RESULTS

### 3.1 Comparing Lowest-Risk Categories: Zero-ABDR Antigen Mismatch and Zero-DR/DQ Eplet Mismatch

Figure 1 shows the percentage of well-matched donors at the antigen (aMM) or eplet (epMM) mismatch level for candidates of each racial/ethnic group. The average percentage of zero-ABDR aMM donors was 0.09% for White candidates, 0.01% for Asian candidates, 0.02% for Black candidates, and 0.03% for Hispanic/Latino candidates. The average percentage of zero-DR/DQ epMM donors was 1.42% for White candidates, 0.43% for Asian candidates, 0.87% for Black % for Hispanic/Latino candidates.

**Figure 1.**
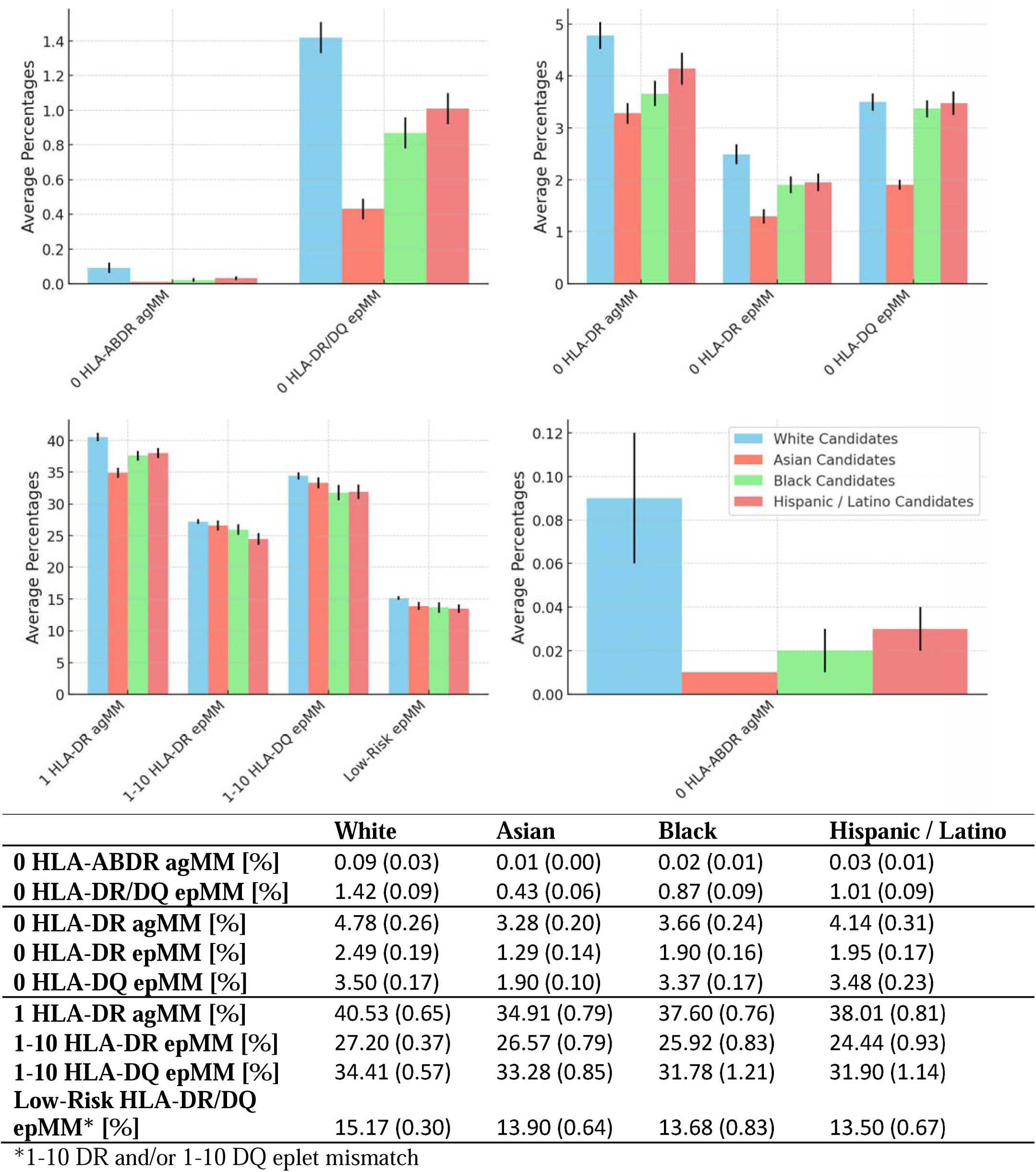
Expected percentages of well-matched donors by candidate ethnicity for various low risk antigen and eplet mismatch categories. Average percentages (and standard deviation) summarize the results across 20 replicate simulated donor pools.

Figure 2 illustrates the expected relative percentage of well-matched donors for White candidates versus Asian, Black, or Hispanic/Latino candidates across 20 simulated donor pools, each with the ethnic composition of OPTN deceased donors. The average percentage of zero-ABDR aMM donors for White candidates was 9.86 times higher than for Asian, 4.97 times higher than for Black, and 3.21 times higher than for Hispanic/Latino candidates. The average percentage zero-DR/DQ epMM donors for White candidates was 3.26 times higher than for Asian, 1.63 times higher than for Black, and 1.41 times higher than for Hispanic/Latino candidates. Comparing the percentage of well-matched donors for zero-ABDR antigen versus zero-DR/DQ epMM, eplet matching was 3.01 times less racially disparate for Asian, 3.05 times less racially disparate for Black, and 2.21 times less racially disparate for Hispanic/Latino candidates compared to antigen-level matching. The zero-DR/DQ epMM risk category significantly increased the percentage of well-matched donors compared to zero-ABDR aMM by 16 times for White, 43 times for Asian, 45 times for Black, and 34 times for Hispanic/Latino candidates.

**Figure 2.**
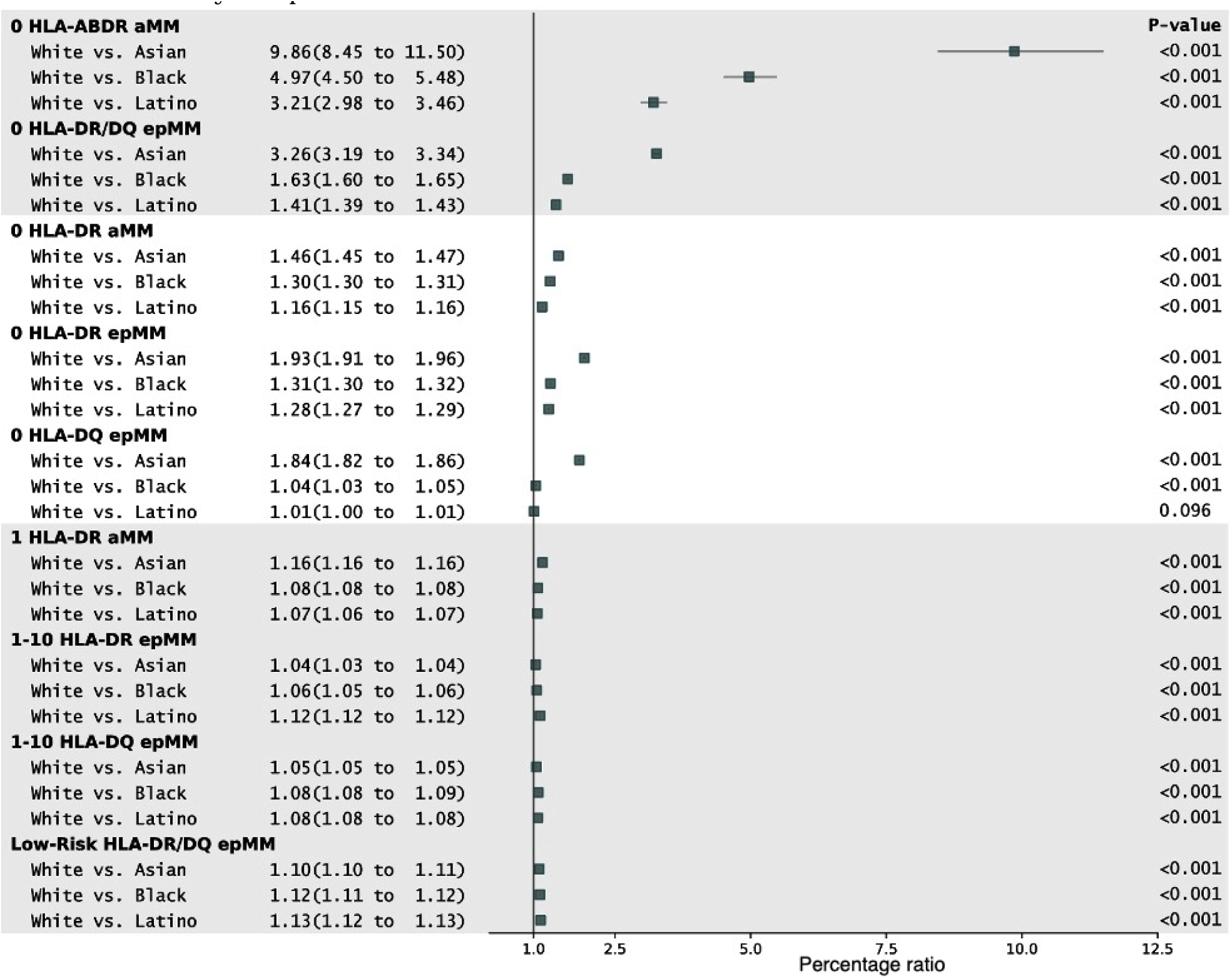
Ethnic disparity metrics for access to well-matched donors. Ratios of average percentage of well-matched donors for White candidates relative to the average percentage of well-matched donors for Asian, Black, or Latino candidates, for each of nine antigen and eplet risk categories. The larger the percentage ratio, the greater the ethnic disparity in access to well-matched donors (e.g., White candidates had 9.86 more zero-ABDR aMM donors than Asian candidates). Average point estimates for percentage ratios (and 95% confidence intervals) are provided for each candidate ethnicity comparison.

### 3.3 Comparing zero-DR antigen mismatch, zero-DR eplet mismatch, and zero-DQ eplet mismatch

Among the zero-DR and -DQ risk categories, the zero-DQ epMM was the most equitable category for Black and Latino candidates, whereas zero-DR aMM and zero-DR epMM favored White candidates (Figure 1 and Figure 2). The average percentage of well-matched donors was 1.30 times higher for White than for Black candidates for zero-DR aMM, 1.31 times higher for White than for Black candidates for zero-DR epMM, and only 1.04 times higher for White than for Black candidates for zero-DQ epMM. The average percentage of well-matched donors was 1.16 times higher for White than for Hispanic/Latino candidates for zero-DR aMM, 1.28 times higher for White than for Hispanic/Latino candidates for zero-DR epMM, and only 1.01 times higher for White than for Hispanic/Latino candidates for zero-DQ epMM. Asian candidates were the most disfavored in all risk categories we studied due to having HLA alleles and antigens that are relatively rare in other populations and because Asians compose only a small percentage of deceased donors.

### 3.4 Comparing 1-DR antigen mismatch, 1-10 HLA-DR eplet mismatch, 1-10 HLA-DQ eplet mismatch, and Low-Risk DR/DQ eplet mismatch

At the non-zero mismatch low risk categories, well-matched donors were only 1.04 - 1.16 times more prevalent for White candidates than for ethnic minority candidates, and Low-Risk DR/DQ epMM (1-10 DR and/or 1-10 DQ eplet mismatch) was the most equitable category (Figure 1 and Figure 2). The 1-DR aMM risk category had the largest average percentage of well-matched donors, ranging from 34.9 to 40.5% among racial and ethnic groups (Figure 1). The 1-10 HLA-DR epMM risk category had an average percentage of 24.4-27.2% well-matched donors.

Although without a direct comparator, the 1-10 HLA-DQ epMM risk category was equitable in that the percentage of well-matched donors was similar across candidate ethnicities. There was also a similar percentage of well-matched donors for both the 1 HLA-DQ epMM risk category and the 1 HLA-DR aMM, in a range of 31.8-34.4% (Figure 1). The percentage of well-matched donors in the low-risk DR/DQ epMM risk category ranged between 13.5% and 15.2% among candidate ethnicity.

## 4. DISCUSSION

HLA eplet matching has gained attention as having better stratification of primary allo-immune risk than antigen-level HLA matching.^25,26^ Evidence supporting the deleterious effect of high eplet mismatch load at HLA-DR and -DQ continues to increase. The possibility of redesigning allocation policy to engineer more transplants to be zero or low HLA-DR/DQ eplet mismatch is gaining traction in the transplant community because eplet mismatch is more strongly associated with dnDSA formation and graft failure than antigen-level mismatch. However, because prioritizing antigen-level mismatch historically created ethnic disparities in kidney allocation, we designed a simulation study using allele-level HLA genotyping data to investigate whether prioritizing eplet mismatch would avoid such disparities. We calculated the percentage of well-matched donors for different ethnicities in replicate donor pools, each with a similar ethnic composition to the OPTN deceased donor pool. To measure ethnic disparity, we calculated the relative percentage of well-matched donors for Asian, Black, Hispanic/Latino, and White candidates using nine different matching risk categories, comparing HLA antigen-level and HLA eplet-level matching. Our results indicate that giving priority for HLA-DR and/or HLA-DQ eplet mismatch would not increase ethnic disparities and could decrease them substantially compared to zero-ABDR antigen mismatch, which is now granted high priority for deceased donor kidney allocation.

In the racially and ethnically imbalanced US deceased donor pool, we showed that non-White candidates are up to 10-times less likely to find the lowest-risk category of zero-ABDR antigen mismatch donor than White candidates. Relative to zero-ABDR antigen mismatch, the lowest-risk zero-DR/DQ eplet mismatch category substantially reduces ethnic disparities in matching likelihoods, i.e., by 3-fold for Black and Asian and by 2-fold for Hispanic/Latino candidates. The zero-DR/DQ eplet risk category also increases the percentage of well-matched donors, especially for racial and ethnic minority candidates, offering them 37-51 times more well-matched donors compared with zero-ABDR antigen mismatch. Among the remaining lower risk categories, 1 HLA-DR aMM, 1-10 HLA-DR epMM, and 1-10 HLA-DQ epMM categories nearly erased racial and ethnic disparities.

For Black and Hispanic/Latino populations, the most equitable risk category was zero-DQ epMM, eliminating the disparities almost completely. Tran et al.^17^ found the HLA-DQ eplets to be the most shared among a heterogenous pool of 2000 kidney donors and recipients, which supports our finding that zero HLA-DQ epMM has the potential to be the most equitable risk stratification method. With an increasing role for HLA-DQ and HLA-DR matching in kidney transplantation and the weight of evidence supporting the deleterious effect of HLA-DQ and HLA-DR dnDSA and graft rejection,^11,25,27^ prioritizing zero-DQ epMM and/or zero-DR epMM donors would reduce allo-immune risk without increasing racial and ethnic disparities. Tambur et al.^28^ found that mismatches at HLA-DQ are not only correlated with rejection but that dnDSA targeting donor HLA-DQ antigens are the most common antibodies post-transplant.

As compared to antigen mismatch, eplet mismatch analysis is a more precise method for primary allo-immune risk assessment. Wiebe et. al. found that a load of >10 HLA-DR and HLA-DQ mismatched eplets is a strong predictive biomarker for the development of HLA-DR and -DQ dnDSA (AUC 0.72 for HLA-DR and DQ), outperforming traditional HLA-DR/DQ antigen mismatch (AUC 0.54 for HLA-DR and 0.58 for HLA-DQ).^25^ Sapir-Pichhadze and colleagues clearly demonstrated a significant correlation between the number of mismatched HLA-DR and -DQ eplet and the likelihood of graft failure in an imputed SRTR dataset.^12^ For every ten mismatched HLA-DR and -DQ eplets, the hazard ratio was 1.35 and 1.29, respectively, with 95% confidence intervals ranging from 1.01 to 1.81 and 1.01 to 1.67 (p = 0.05). Since eplet mismatch is a superior indicator of HLA compatibility, the current prioritization of 1 HLA-DR aMM overestimates the number of donors that are truly low primary allo-immune risk. In our simulation, 1 HLA-DR aMM identified an average of 35.0-40.5% matched donors among racial and ethnic groups and improved disparities. However, only 24.4-27.2% of donors in the pool were low-risk HLA-DR epMM, 31.8-34.4% were low-risk HLA-DQ epMM, and 13.5-15.2% were low-risk HLA-DR/DQ epMM donors. The donors who are actually low-risk can be more accurately identified by the eplet mismatch approach.

Prioritizing eplet mismatch in deceased donor kidney allocation would require implementation of rapid deceased donor allele-level genotyping in clinical labs, however the methods are still under evaluation. A key advantage of our study is that it utilized allele-level HLA genotypes determined by next-generation sequencing in the setting of organ transplantation. Continuing research in HLA mismatch and outcomes to build a strong evidence base to support policy changes is also necessary. Tambur et al.^29^ provided a comprehensive commentary on issues to address before using eplet mismatch in organ allocation.

There are several limitations in our study. Although we resampled the donor population to match the racial/ethnic makeup of the OPTN deceased donor pool, racial/ethnic minorities are represented the NKR dataset (Figure 1). We looked at the impact of eplet matching only on a crude aggregation into four broadly-defined self-identified racial and ethnic groups (Asian, Black, Latino, and White), which conceals population substructure and heterogeneity in match likelihoods for more detailed subpopulations. However, our simulation results are consistent with registry data analysis^8^ showing that Asian, Black, and Latino candidates are less likely to find a zero-ABDR antigen mismatch donor. We considered eplet mismatch risk categories only as the sum of eplet mismatches at a given locus. Future studies could investigate the impact of other risk categories on equity considering single molecule eplet mismatch, surface-exposed mismatched amino-acid (HLA-EMMA) and by the amount of peptides derived from HLA mismatched donor proteins that are indirectly presented by recipient Class II molecules to CD4+ T cells (PIRCHE-II). Although Wiebe et.al. have demonstrated that their different B cell molecular mismatch paradigms express high degree of correlation (r^2^ = 0.85-0.96),^30^ these mismatch methods must be verified in a much more heterogeneous population, possibly leading to alternative optimal mismatch thresholds for risk stratification. Since our cohort comes from the living donation program, each potential recipient might be related to at most one donor, which could slightly inflate the likelihood of finding a well-matched donor.

Our study provides a new level of evidence that compared to the currently prioritized antigen mismatch risk categories, eplet matching would not increase and might decrease ethnic and racial disparities for lowest-risk categories, and eplet matching identifies more truly low allo-immune risk donors. Eplet immunogenicity studies and long-term outcomes research on low allo-immune risk donors remains necessary to refine optimal risk stratification paradigms and improve understanding of the immunologic mechanisms involved. However, eplet mismatch clearly stratifies risk better than antigen mismatch and our data supports eplet mismatch could be incorporated into kidney deceased and living donor allocation algorithms in the near future without worsening racial and ethnic disparities. The impact of eplet mismatch should next be evaluated more rigorously using full simulated allocation models, which would require integration of allele-level HLA genotyping data into allocation modeling software.

## Data Availability

Data can be requested from the NKR.

## Acknowledgments

We thank Matt Ronin and the NKR Team for providing the study population and the eplet mismatch load at HLA-ABC, HLA-DR, HLA-DQ, and HLA-DP.

## ABBREVIATIONS

ASTS: American Society of Transplant Surgeons
ABMR: Antibody Mediated Rejection
agMM: HLA Antigen Mismatch
ASTS: American Society of Transplant Surgeons
dnDSA: *de-novo* Donor Specific Antibody
epMM: HLA Eplet Mismatch
NGS: Next Generation Sequencing
NKR: National Kidney Registry
OPTN: Organ Procurement & Transplantation Network
SRTR: Scientific Registry for Transplant Recipients
STAR: Sensitization in Transplantation: Assessment of Risk
TCMR: T cell Mediated Rejection
UNOS: United Network for Organ Sharing

## REFERENCES

1. Williams RC, Opelz G, McGarvey CJ, Weil EJ, Chakkera HA. The Risk of Transplant Failure With HLA Mismatch in First Adult Kidney Allografts From Deceased Donors. Transplantation. 2016;100(5):1094–1102.

2. Foster BJ, Dahhou M, Zhang X, Platt RW, Smith JM, Hanley JA. Impact of HLA mismatch at first kidney transplant on lifetime with graft function in young recipients. Am J Transplant. 2014;14(4):876–885.

3. Ashby VB, Port FK, Wolfe RA, et al. Transplanting kidneys without points for HLA-B matching: consequences of the policy change. Am J Transplant. 2011;11(8):1712–1718.

4. Roberts JP, Wolfe RA, Bragg-Gresham JL, et al. Effect of changing the priority for HLA matching on the rates and outcomes of kidney transplantation in minority groups. N Engl J Med. 2004;350(6):545–551.

5. Hall EC, Massie AB, James NT, et al. Effect of eliminating priority points for HLA-B matching on racial disparities in kidney transplant rates. Am J Kidney Dis. 2011;58(5):813–816.

6. Robinson A, Lindblad K, Stewart D, et al. Racial Differences in HLA Mismatch Potential Among Kidney Registrations [abstract]. Am J Transplant. 2022;(22 (suppl 3)):3–5.

7. ASTS Responses to OPTN Proposals Open for Public Comment. Published 2022. https://unos.my.salesforce.com/sfc/p/#80000000bHuz/a/3n000000qBSO/8ORMmdsyvTXLPRYq5PUrjPccTUzYvbITaPOIt6m4tlM

8. Tambur AR, Bestard O, Campbell P, et al. Sensitization in transplantation: Assessment of Risk 2022 Working Group Meeting Report. Am J Transplant. 2023;23(1):133–149.

9. Huang Y, Dinh A, Heron S, et al. Assessing the utilization of high-resolution 2-field HLA typing in solid organ transplantation. Am J Transplant. 2019;19(7):1955–1963.

10. De Santis D, Truong L, Martinez P, D’Orsogna L. Rapid high-resolution HLA genotyping by MinION Oxford nanopore sequencing for deceased donor organ allocation. HLA. 2020;96(2):141–162.

11. Wiebe C, Pochinco D, Blydt-Hansen TD, et al. Class II HLA epitope matching-A strategy to minimize de novo donor-specific antibody development and improve outcomes. Am J Transplant. 2013;13(12):3114–3122.

12. Sapir-Pichhadze R, Zhang X, Ferradji A, et al. Epitopes as characterized by antibody-verified eplet mismatches determine risk of kidney transplant loss. Kidney Int. 2020;97(4):778–785.

13. Kosmoliaptsis V, Mallon DH, Chen Y, Bolton EM, Bradley JA, Taylor CJ. Alloantibody Responses After Renal Transplant Failure Can Be Better Predicted by Donor-Recipient HLA Amino Acid Sequence and Physicochemical Disparities Than Conventional HLA Matching. Am J Transplant. 2016;16(7):2139–2147.

14. Bestard O, Meneghini M, Crespo E, et al. Preformed T cell alloimmunity and HLA eplet mismatch to guide immunosuppression minimization with tacrolimus monotherapy in kidney transplantation: Results of the CELLIMIN trial. Am J Transplant. 2021;21(8):2833–2845.

15. Philogene MC, Amin A, Zhou S, et al. Correction to: Eplet mismatch analysis and allograft outcome across racially diverse groups in a pediatric transplant cohort: a single-center analysis. Pediatr Nephrol. 2020;35(4):719.

16. Wiebe C, Nickerson PW. Role of HLA molecular mismatch in clinical practice. Hum Immunol. 2022;83(3):219–224.

17. Tran JN, Günther OP, Sherwood KR, et al. High-throughput sequencing defines donor and recipient HLA B-cell epitope frequencies for prospective matching in transplantation. Commun Biol. 2021;4(1):583.

18. Niemann M, Lachmann N, Geneugelijk K, Spierings E. Computational Eurotransplant kidney allocation simulations demonstrate the feasibility and benefit of T-cell epitope matching. PLoS Comput Biol. 2021;17(7):e1009248.

19. Bekbolsynov D, Mierzejewska B, Khuder S, et al. Improving Access to HLA-Matched Kidney Transplants for African American Patients. Front Immunol. 2022;13(March):832488.

20. National Kidney Registry - Medical Board Policies. Accessed June 1, 2023. https://www.kidneyregistry.org/for-centers/medical-board-policies/

21. Kausman JY, Walker AM, Cantwell LS, Quinlan C, Sypek MP, Ierino FL. Application of an epitope-based allocation system in pediatric kidney transplantation. Pediatr Transplant. 2016;20(7):931–938.

22. Kaur N, Kransdorf EP, Pando MJ, et al. Mapping molecular HLA typing data to UNOS antigen equivalents. Hum Immunol. 2018;79(11):781–789.

23. Taoti Enterprises International, Web Design, Marketing. - national Kidney Registry - facilitating living donor transplants. Accessed June 13, 2024. https://portal.kidneyregistry.org/hla-tools/epitope-mismatch

24. Bezstarosti S, Bakker KH, Kramer CSM, et al. A comprehensive evaluation of the antibody-verified status of eplets listed in the HLA Epitope Registry. Front Immunol. 2021;12:800946.

25. Wiebe C, Kosmoliaptsis V, Pochinco D, et al. HLA-DR/DQ molecular mismatch: A prognostic biomarker for primary alloimmunity. Am J Transplant. 2019;19(6):1708–1719.

26. Davis S, Wiebe C, Campbell K, et al. Adequate tacrolimus exposure modulates the impact of HLA class II molecular mismatch: a validation study in an American cohort. Am J Transplant. 2021;21(1):322–328.

27. Senev A, Coemans M, Lerut E, et al. Eplet Mismatch Load and De Novo Occurrence of Donor-Specific Anti-HLA Antibodies, Rejection, and Graft Failure after Kidney Transplantation: An Observational Cohort Study. J Am Soc Nephrol. 2020;31(9):2193–2204.

28. Tambur AR, Kosmoliaptsis V, Claas FHJ, Mannon RB, Nickerson P, Naesens M. Significance of HLA-DQ in kidney transplantation: time to reevaluate human leukocyte antigen–matching priorities to improve transplant outcomes? An expert review and recommendations. Kidney Int. 2021;100(5):1012–1022.

29. Tambur AR, Das R. Can We Use Eplets (or Molecular) Mismatch Load Analysis to Improve Organ Allocation? The Hope and the Hype. Transplantation. 2023;107(3):605–615.

30. Wiebe C, Kosmoliaptsis V, Pochinco D, Taylor CJ, Nickerson P. A Comparison of HLA Molecular Mismatch Methods to Determine HLA Immunogenicity. Transplantation. 2018;102(8):1338–1343.

